# Opioid Use Stigmatization and Destigmatization in Healthcare Professional Social Media

**DOI:** 10.1101/2021.10.19.21265210

**Authors:** S. Scott Graham, Fiona N. Conway, Richard Bottner, Kasey Claborn

## Abstract

Stigmatization of opioid use constitutes a significant barrier to addressing the opioid crisis. Increasing use of social media by healthcare professionals provides an opportunity to foster destigmatization. However, little is known about stigmatization and destigmatization within healthcare professional social media communities. Accordingly, this study investigates the use of stigmatizing and destigmatizing language in three such communities: Medical Twitter, Public Health Twitter, and Epidemiology Twitter. Using a dataset of 2,319 tweets discussing opioids and associated with these Twitter communities, we analyzed each tweet for evidence of stigmatizing or destigmatizing language based on guidance from the National Institute on Drug Abuse. The results indicate that overall use of both stigmatizing and destigmatizing language is currently low across communities compared to the overall volume of opioid-related content. Additionally, there are measurable changes in stigmatizing and destigmatizing language on quarterly bases between 2012 and 2020. During this time, Public Health Twitter has seen a quarterly 19% reduction in rates of stigmatizing (IRR = 0.81, 95% CI 0.67 to 0.97), and all communities have experienced a quarterly 57% increase in destigmatizing language (IRR = 1.57, 95% CI: 1.33 to 1.85). This study also reveals that tweets containing stigmatizing and destigmatizing language receive minimal user engagement (measured by likes, retweets, quote tweets, and comments). While the longitudinal findings on increasing use of destigmatizing language are promising, they also indicate a need for increased efforts to encourage broader use of destigmatizing language. Leveraging the social learning potentials of Twitter offers one promising pathway for future initiatives.

## Introduction

The opioid epidemic continues to be a significant challenge for individuals, communities, and healthcare systems. Preliminary data show that overdose deaths in the United States rose 29.4% in 2020 to an estimated 93,331, including 69,710 involving opioids (Ahmad, Rossen, Sutton, 2021). This is the latest in a concerning trend that includes a 1,040% increase in synthetic opioid overdosed since 2013 (Mattheson, et al., 2021). Emerging evidence also indicates that the opioid epidemic may be exacerbated by COVID-19 (Linas, et al., 2021). While overall emergency department visits declined between March and September 2020, there was an increase in visits for overdose including opioid-related overdose (Holland, et al., 2021). In addition to health effects and loss of life, the opioid epidemic has also been linked to significant economic impacts. The US Society of Actuaries estimates that non-medical use of opioids resulted in $21.9 billion in productivity losses (Davenport, Weaver, and Caverly, 2019). And the overall economic impact of the opioid epidemic in the US was estimated at $1.02 trillion for 2017 (Luo, Li, and Florence, 2021). Given the wide-ranging deleterious effects of the opioid epidemic, it is critical that healthcare professionals actively investigate new approaches to attend to this crisis.

While there are many challenges that blunt efforts to effectively address the opioid epidemic, stigma experienced by people who use opioids is one of the greatest barriers (Wakeman & Rich, 2019; McGinty & Barry, 2020; Tsai, Kiang, Barnett, et al., 2019). Stigmatization results in delays seeking care and premature treatment termination (Hammarlund, Crapanzo, Luce, Mulligan, & Ward, 2018; Luoma, 2010). Despite the ready availability of research on the dangers of opioid use stigmatization, stigmatizing language continues to prevail in public, private, and clinical contexts (Hammarlund, Crapanzo, Luce, Mulligan, & Ward, 2018; Luoma, 2010). Using stigmatizing language is learned behavior. As such, efforts to change this behavior require exploring the mechanisms that underlie behavioral and social learning. Early investigations by psychologists Thorndike (1905), Watson (1924), Pavlov (1927), and Skinner (1938) focused on the theory of conditioning in behavioral learning. Conditioning, often referred to as learning by association (Stangor & Walinga, 2014), centers on the relationship between stimuli and response. Watson’s (1924) and Pavlov’s (1927) work focused on classical conditioning—the repeated pairing of two stimuli to provoke a learned response. For example, repeated pairing of seeing a toy car with feeling electric shock results in the learned behavior of avoiding toy cars. Classical conditioning focuses on the learning that occurs *before* the behavior is initiated. In contrast, Thorndike’s (1905) and Skinner’s (1938) work explored operant conditioning—the process of learning through punishment or reward *after* behavior has occurred. The learned association occurs *after* the behavior by pairing it and its consequence (Tukel, 2020). For example, if a child hits someone and is praised for their behavior, hitting others is associated with a positive outcome, and the behavior is reinforced. Whereas, if the child is punished for hitting others, the behavior is associated with a negative outcome and therefore is discouraged. Learning by association through operant conditioning provides a solid theoretical explanation for the continued use of stigmatizing language regarding people who use opioids.

The use of stigmatizing language regarding this population rarely results in censure (i.e., punishment). In some instances, people who use stigmatizing language gain rewards, such as being elevated to the status of a professional “expert” on issues regarding substance use, despite the known damage associated with such language. Another important theory that adds further insight regarding the entrenchment of the use of stigmatizing language is Bandura’s social learning (1977). Bandura’s theory, grounded in classical conditioning and operant conditioning principles, explains how we learn through social experiences. He expands on the conditioning theories by positing that direct experience is not requisite for behavioral learning. He argues that learning also occurs through observing the behavior of models (i.e., other people), evaluating the reward or punishment models experience, and choosing to imitate the behaviors or not. In language related to people who use opioids, individuals observe their models (e.g., journalists, experts, treatment providers, etc.) using stigmatizing language, judge whether the behavior is censured or encouraged, and may then choose to imitate the behavior.

Given the documented effects of stigma on treatment and recovery, there have been increasing calls to embrace stigma reduction strategies in public, private, and clinical contexts (McGinty & Barry, 2020; Howard, 2015). Recommended stigma reduction strategies include the replacement of stigmatizing language with person-first language (Atayde, Hauc, Bessette, Danckers, & Saitz, 2021; Kelly & Westerhoff, 2010; Soloner, McGinty, Beletsky, Bluthenal, Beyrer, Botticelli, & Sherman, 2018; Ashford, Brown, & Curtis, 2019) and solution-focused language (McGinty, Goldman, Pescosolido & Barry, 2015). Additionally, emerging research indicates that the strategic use of positive drug stories (Engell, Bright, Barrett, & Allen, 2020) and sympathetic narratives (Kennedy-Hendricks, McGinty, & Barry, 2016; Heley, Kennedy-Hendricks, Niederdeppe, Barry, 2019) can lead to stigma reduction. Given the power that destigmatizing language can have to improve outcomes for people who use opioids, it is essential that healthcare providers and public health professionals pursue destigmatizing efforts both within and outside of clinical settings. Due to its potential for expansive influence, social media provides a novel opportunity for healthcare professionals to leverage the principles of operant conditioning and social learning theories to engage in stigma reduction at scale. Specifically, social media has provided an unpreceded opportunity for the public to access and engage with healthcare professional communication. Increasing numbers of healthcare professionals are using social media, not just in personal capacities but also in professional and para-profession roles (Kantor, Bright & Burtchell, 2018). This phenomenon has recently accelerated in the wake of the COVID-19 pandemic. In the early days of the pandemic, a public desperate for emerging information turned to healthcare provider social media accounts for up-to-date information (Graham, 2020; Hu-Undark, December 2, 2020). As the public sought information, Twitter, for example, rushed to provide public health experts and clinicians with the blue checkmark of “verified” status. The goal here was to help the public connect with “trusted” experts (Neporent, 2020). Of course, public access to healthcare professional social media will not guarantee that stigma reduction takes place. There is significant evidence available that indicates use of stigmatizing language is common on popular and social media (McGinty, Kennedy-Hendrics & Barry, 2019; Dekeserdy, Sedney, Razzaq, Haggerty, and Brownstein, 2021). Additionally, research on patient-provider encounters has shown that stigma can be exacerbated by the language healthcare professionals use when talking about opioids and OUD (Stone, Kennedy-Hendricks, Barry, Bachuber, & McGinty, 2021; Dassieu, Heino, Develay, Kaboré, Pagé, Moor et al., 2021). While social media provides an unprecedented opportunity for healthcare professionals to support stigma reduction, the extent to which professionals embrace this opportunity is unknown. Additionally, the extent to which healthcare professionals participate in the more common stigmatizing practices on social media is also unknown.

## Methods

In order to explore this issue, we conducted a study of three healthcare professional social media communities on Twitter, Medical Twitter, Public Health Twitter, and Epidemiology Twitter. Specifically, we sought to investigate the following research questions: 1) What is the prevalence of opioid-related stigmatizing and destigmatizing language in the identified Twitter communities? 2) Has there been a change in the frequency of tweets using stigmatizing or destigmatizing language over time? And 3) How popular are tweets with stigmatizing or destigmatizing language compared to tweets that contain neither stigmatizing nor destigmatizing language? In order to address these questions, we used the Twitter Academic Application Programming Interface (Tornes & Trujillo, January 2021) to collect all tweets that used a relevant hashtag for one of three healthcare professional communities and one or more relevant opioid or OUD-related terms. Opioid and OUD-related search terms were identified based on topical relevance. However, in order to avoid biasing study results, relevant terms that are inherently stigmatizing or stigma reducing were excluded. See Table 1 for a complete list of hashtags and search terms.

**Table 1.**
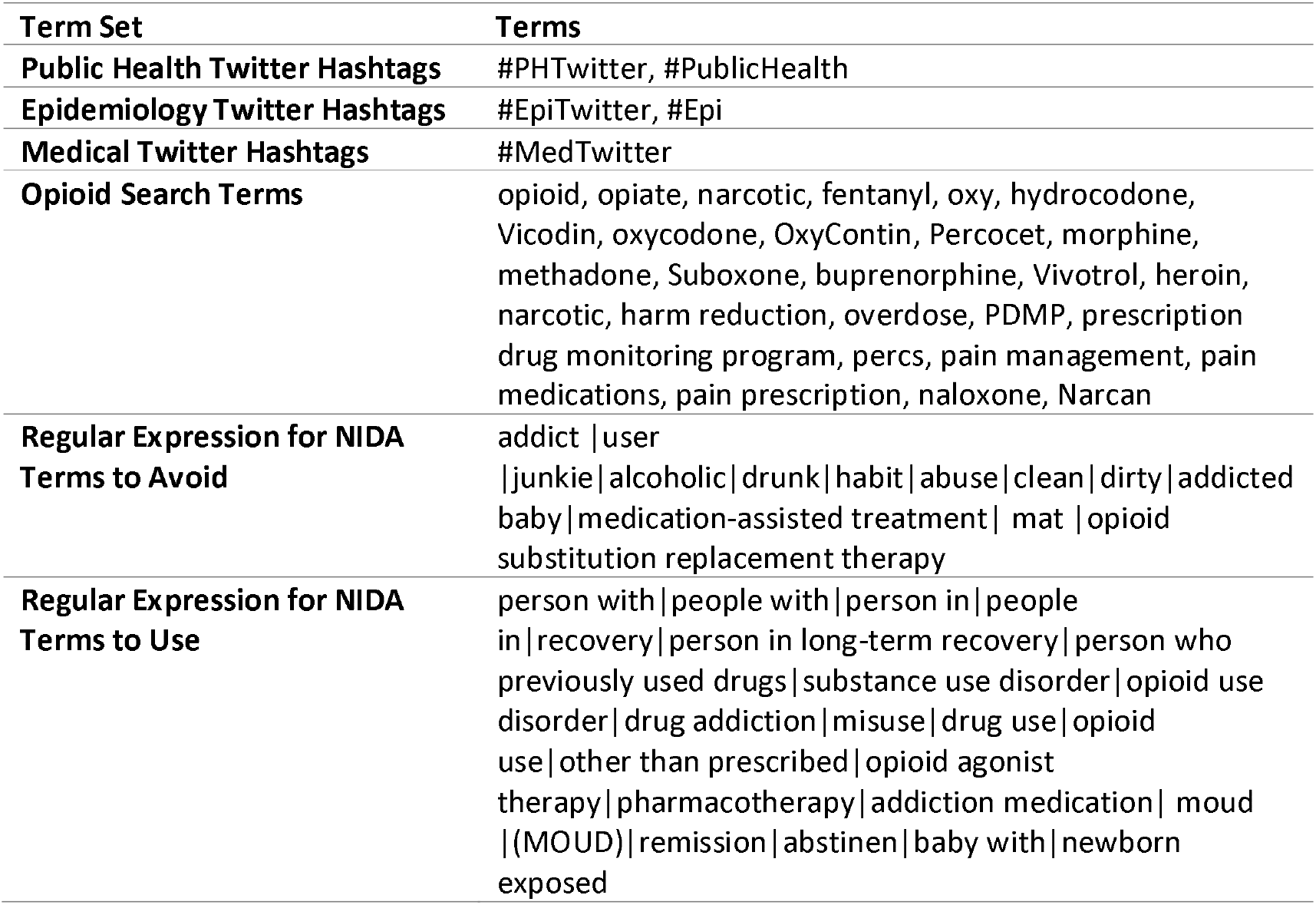
Search Terms for Data Collection and Analysis

**Table 2.**
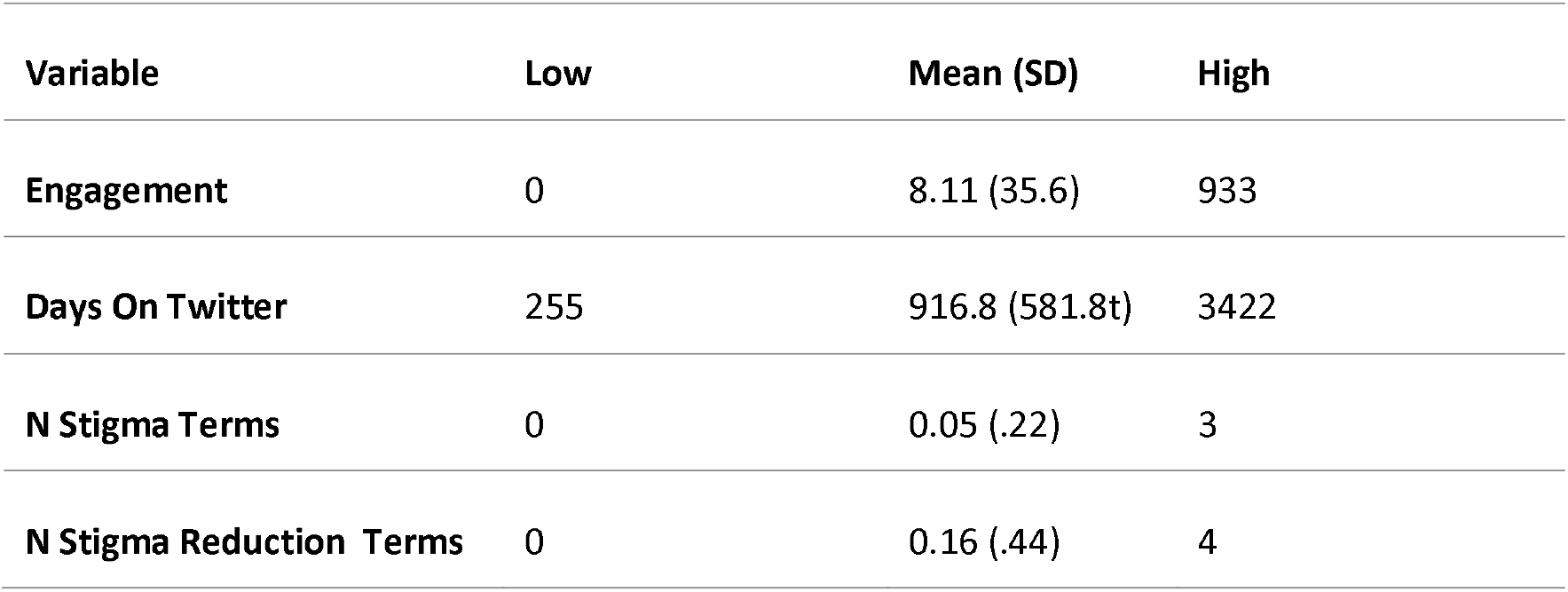
Descriptive Statistics for the Three-Community Twitter Dataset.

| Variable | Low | Mean (SD) | High |
| --- | --- | --- | --- |
| Engagement | 0 | 8.11 (35.6) | 933 |
| Days On Twitter | 255 | 916.8 (581.8t) | 3422 |
| N Stigma Terms | 0 | 0.05 (.22) | 3 |
| N Stigma Reduction Terms | 0 | 0.16 (.44) | 4 |

The Twitter searches retrieved a total of 14,772 tweets posted between 1/1/2012 and 12/31/2020. This included 1,931 tweets from Medical Twitter, 12,529 tweets from Public Health Twitter and 312 tweets from Epidemiology Twitter. In order to have more equal samples for subsequent analysis, we took a random sample of 1,000 tweets from each of the Medical Twitter and Public Health Twitter datasets and 681 tweets that did not contain any of the search terms were removed. Subsequently, the final dataset included a total of 2,319 opioid-related tweets from the three communities.

With the final dataset in hand, we calculated an engagement score for each tweet based on the sum of retweets, quote tweets, likes, and comments. Most tweet engagement occurs in the hours and days after initial posting. Since all tweets collected were at least several months old, time since posting will not bias the engagement scores. We also evaluated each tweet for the presence of stigmatizing or destigmatizing language. We used the lists of “terms to use” and “terms to avoid” published by the National Institute for Drug Abuse (NIDA, 2021) to identify stigmatizating and destigmatizing language related to substance use. We used a regular expressions (regex) framework to operationalize our search for these terms. Table 1 also includes the regex statements used in the analysis.

## Results

All collected tweets were between 255 and 3422 days old with an average age of 916.8. Each tweet contained between 0 and 3 terms to avoid (mean = 0.05) and between 0 and 4 stigma reduction terms (mean = 0.16). Table 1 summarizes these data. Our community-based analysis indicates that the overall prevalence of stigmatizing language is low across healthcare professional Twitter communities. Of the 2,319 tweets with opioid and OUD-related terms 4.49% of Epidemiology, 2.4% of Medical tweets, and 6.4% of Public Health Twitter tweets contained language on NIDA’s list of terms to avoid. In order to determine if there was a significant difference in the prevalence of stigmatizing language by Twitter community, we conducted a three-way community difference between independent proportions test. The three-sample test for equality of proportions found a statistically significant difference between proportions, χ^2^ = 18.975, p<0.0001. Medical Twitter has markedly lower prevalence of tweets using NIDA stigmatizing terms than either Epidemiology Twitter or Public Health Twitter. The overall prevalence of tweets using stigma reduction language is also somewhat low, with 14.1% of Epidemiology Twitter tweets, 14.8% of Medical Twitter postings, and 11.3% of Public Health Twitter tweets containing destigmatizing terms. A three-sample test for equality of proportions found no significant difference between twitter communities, χ^2^ = 5.61, p=0.06052.

We also evaluated the relationships between stigmatizing rate or destigmatizing rate, community affiliation, and time. Quarterly stigma rates and trend lines by community are available in Figures 2a and 2b. Poisson family regression models have proved highly robust for similar time-series data, and given our use of percentage outcomes, a quasi-Poisson framework was most appropriate to this analysis (Kuhn, Davidson, & Durkin, 1994). Within a Poisson family framework, the sample size is sufficient for 95% power even in cases of modest incident rate ratios (IRR) (Singorini, 1991). Therefore, we fit a model to see if date, community membership, or the interaction between the two predicted stigmatization rates in the dataset. For this initial model the only significant predictors were for tweets affiliated with the Public Health Twitter community (p = 0.0242) and for the interaction between the Public Health Twitter community and the date (p = 0.0243). Subsequently, we fit a new model evaluating the relationship between the data and the stigma language rates using only the Public Health Twitter data. The posting quarter was a significant predictor (p = 0.0252). The results indicate that each successive quarter is associated with 19% reduction in the rate of stigmatizing language (IRR = 0.81, 95% CI 0.67 to 0.97). In order to evaluate potential changes in NIDA recommended language usage, we fit another quasi-Poisson model to test date and community as predictors for stigmatization rates. Date was the only significant predictor (p = 0.000111). Subsequently, we dropped community from the model. Date remained significant (p = 1.07e-06), and with each passing quarter we see a 57% increase in destigmatizing language (IRR = 1.57, 95% CI: 1.33 to 1.85).

**Figure 1.**
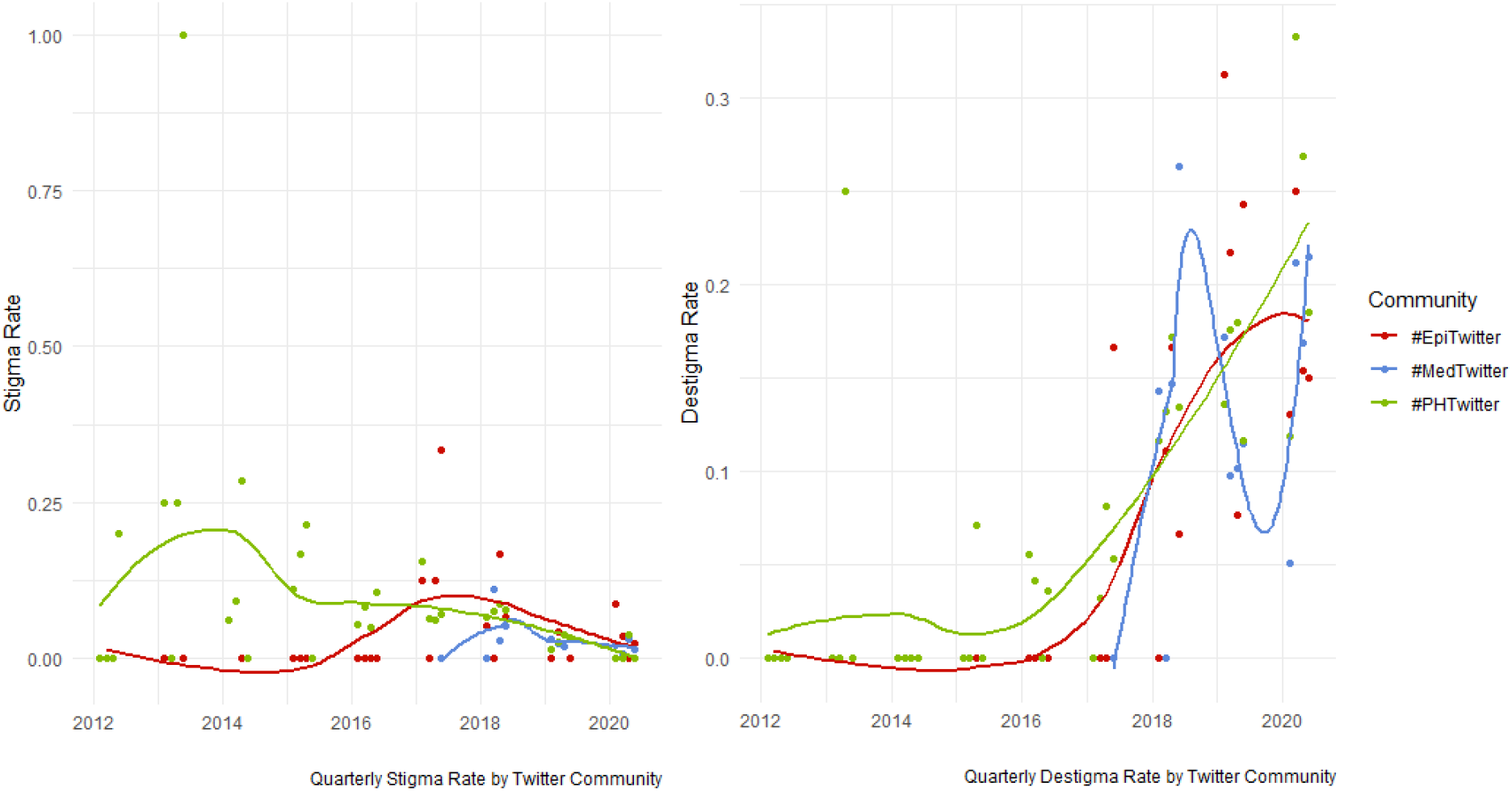
Trends in Quarterly stigmatizing language (A) and stigma reduction (B) rates by Twitter community.

**Figure 2.**
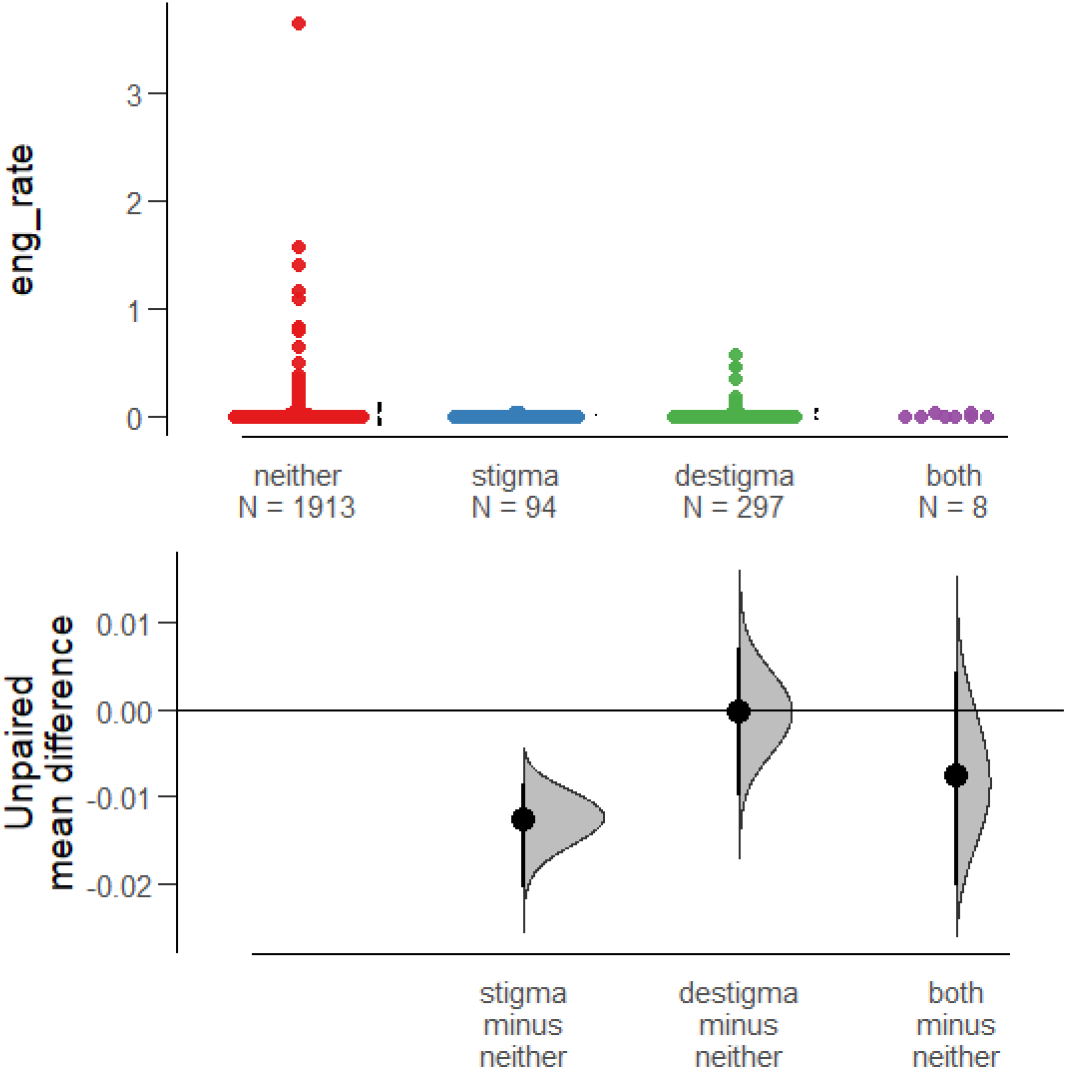
Mean difference estimation for tweets with stigmatizing language, destigmatizing language, or both when compared to tweets that contained neither.

The vast majority of the opioid-related tweets in selected Twitter communities do not use either stigmatizing or destigmatizing language. Therefore, we sought to evaluate if tweets with stigmatizing or destigmatizing language resulted in different user engagement profiles when compared to the majority of tweets. That is, do tweets with stigmatizing or destigmatizing language result in markedly different rates of likes, retweets, quote tweets, or comments? Average engagement per tweet in the dataset is low (8.11 interactions) as is the case for most tweets broadly. Additionally, the range is large (0 to 933 interactions). Thus, to evaluate differential engagement with stigmatizing or destigmatizing content, we used bias-corrected and accelerated (BCa) mean difference estimation from the estimation statics framework (Ho, Tumkaya, Aryal, Choi, & Claridge-Chang, 2019). The results indicate that, on average, tweets containing stigmatizing language received lower rates of engagement (retweets, comments, likes, and quote tweets) than tweets that contained stigmatizing or destigmatizing language (mean difference = −4.77, 95% CI: −7.36 to −2.8). The analysis also shows that there is no meaningful difference in average user engagement with tweets that contain destigmatizing language are compared to tweets that contain neither stigmatizing nor destigmatizing language (mean difference = −0.04, 95% CI: −3.34 to 2.79). The results are similar for the few tweets that contain both stigmatizing and destigmatizing language (mean difference = −1.82, 95% CI −8.71 to 8.38). Figure 2 details these findings.

## Discussion

Broadly, the results presented here indicate that members of the Medical Twitter, Epidemiology Twitter, and Public Health Twitter communities do not use high rates of stigmatizing language. Interesting, Medical Twitter has the lowest rates of stigmatizing language among communities. While this study cannot definitively identify why this might be the case, the rise in stigma reduction continuing medical education credits (which are not often required among non-medical epidemiology and public health professionals) may help explain the differential. Furthermore, the available data indicate that in Public Health Twitter, there has been a marked reduction in the use of stigmatizing language between 2012 and 2020. The Medical Twitter and Epidemiology Twitter communities have not been as active for as long on matters of opioid use, and therefore there is insufficient longitudinal data to identify a change in stigmatizing language use in these communities. The observed changes coincide broadly with increasing academic concerns about the effects of stigmatization and public health efforts designed to address the broad use of stigmatizing language. Nevertheless, the observed trend in Public Health Twitter’s decreasing use of stigmatizing language accelerates between 2014 and 2015, approximately the time that several well-cited papers on stigmatization were published (Barr, McGinty, Pescosolido, & Goldman, 2014; Kelley, Wakeman, Saitz, 2015). Importantly, tweets using stigmatizing language are generally not well rewarded with engagement in the form of likes, comments, retweets, or quote tweets.

Despite these optimistic findings about low and decreasing use of stigmatizing language on Twitter, person-first and other stigma reducing terms are not well represented across healthcare professional Twitter communities. Only about 11-15% of discussions about opioids make use of stigma reducing language. Given that use of stigma reduction language is not well rewarded in terms of engagement, it is, perhaps, unsurprising that it is such a small part of overall opioid discussions within healthcare professional Twitter communities. Fortunately, the usage rate for these terms appears to be increasing quickly. While usage is largely flat between 2012 and 2016, the adoption of stigma reducing language appears to have accelerated between 2016 and 2017. These changes, too, coincide with increasing visibility of stigma reduction strategies and scholarly publication in these areas.

## Conclusion

Social marketing has previously been identified as a potentially effective way of encouraging destigmatization of opioid use (Lavack, 2007). Given the broad access the public now has to healthcare social media, healthcare professionals have an unprecedented opportunity to engage in destigmatizing social marketing at scale. However, the findings presented in this article suggest that many healthcare professionals are not taking full advantage of this opportunity. While rates of stigmatizing language are low, destigmatizing language use rates are also low compared to the overall volume of opioid-related content. While use of destigmatizing language on Twitter has increased in recent quarters, it is still generally less than 20% of opioid-related content in Medical, Public Health and Epidemiology Twitter communities. Nevertheless, observed changes in stigmatizing and destigmatizing language use indicate that positive change is possible. In addition to increased use of destigmatizing language, healthcare professionals who use Twitter might also consider engaging in additional efforts to support broader use of destigmatizing language. Content and behavior on social media are functions of both social learning and broader social-media-specific social norms (Dehghani, et al., 2016). Both observation of prior user behavior and operant conditioning deployed through engagement (likes, retweets, quote tweets, and comments) on social networks can affect post frequency and user content (Dehghani, et al., 2016, Brady, Crockett and Van Bavel, 2020). However, the most recent evidence indicates that social learning is especially powerful in shaping social media content. A recent evaluation of moral outrage on Twitter found not only that reinforcement learning is more predictive of future content, but also that positive user engagement is more salient than a lack of user engagement (Brady, McLoughlin, Doan, and Crockett, 2021). These analyses suggest that while use of destigmatizing language is increasing in selected Twitter communities, increases could be accelerated with additional attention to reinforcement learning strategies. That is, if members of Medical, Public Health, or Epidemiology Twitter communities more frequently celebrated the presence of destigmatizing language, we should expect to see a corresponding increase in the use of that language.

Future research could support efforts in these areas through (1) developing more expansive approaches to identifying stigmatization and destigmatization on social media and (2) evaluating which modes of user engagement most effectively encourage broader adoption of destigmatization techniques. While the NIDA list of terms to use and terms to avoid is an excellent starting point for destigmatization research and practice, the available literature indicates that a broader array of communication strategies (beyond word choice) can effectively support destigmatization. Three promising strategies include a focus on solutions over causes (McGinty, Goldman, Pescosolido & Barry, 2015), positive drug stories (Engell, Bright, Barrett, and Allen, 2020), and sympathetic narratives (Kennedy-Hendricks, McGinty, & Barry, 2016; Heley, Kennedy-Hendricks, Niederdeppe, Barry, 2019). The development of new techniques and approaches designed to identify these strategies at scale on social media can help researchers better understand and track destigmatization efforts online. Additionally, the available research on social media moral outrage indicates that certain forms of user engagement are more effective than others in shifting social media content (Dehghani, et al., 2016, Brady, Crockett and Van Bavel, 2020). Additional research on social media destigmatization should evaluate if certain modes of engagement or if certain educational techniques deployed within comments or quote tweets are more likely to encourage future use of destigmatizing language.

## Data Availability

All data produced in the present study are available upon reasonable request to the authors.

## References

Ahmad, F.B., Rossen, L.M., & Sutton, P. (2021). Provisional drug overdose death counts. National Center for Health Statistics. https://www.cdc.gov/nchs/nvss/vsrr/drug-overdose-data.htm.

Ashford, R. D., Brown, A., & Curtis, B. (2019). Expanding language choices to reduce stigma: A Delphi study of positive and negative terms in substance use and recovery. Health Education, 119(1), 51–62.

Atayde, A.M.P., Hauc, S.C., Bessette, L.G., Danckers, H. & Saitz, R. (2021) Changing the narrative: A call to end stigmatizing terminology related to substance use disorders, Addiction Research & Theory, 29:5, 359–362,

Bandura, A. (1977). Social learning theory. Englewood Cliffs, NJ: Prentice Hall.

Barry, C. L., McGinty, E. E., Pescosolido, B. A., & Goldman, H. H. (2014). Stigma, discrimination, treatment effectiveness, and policy: Public views about drug addiction and mental illness. Psychiatric Services, 65(10), 1269–1272.

Brady, W. J., Crockett, M. J., & Van Bavel, J. J. (2020). The MAD model of moral contagion: The role of motivation, attention, and design in the spread of moralized content online. Perspectives on Psychological Science, 15(4), 978–1010.

Brady, W. J., McLoughlin, K., Doan, T. N., & Crockett, M. (2021). How social learning amplifies moral outrage expression in online social networks. Science Advances, 7(33), eabe5641.

Dassieu, L., Heino, A., Develay, É., Kaboré, J. L., Pagé, M. G., Moor, G., … & Choinière, M. (2021). “They think you’re trying to get the drug”: Qualitative investigation of chronic pain patients’ health care experiences during the opioid overdose epidemic in Canada. Canadian Journal of Pain, 5(1), 66–80.

Dehghani, M., Johnson, K., Hoover, J., Sagi, E., Garten, J., Parmar, N. J., … & Graham, J. (2016). Purity homophily in social networks. Journal of Experimental Psychology: General, 145(3), 366.

Dekeseredy, P., Sedney, C. L., Razzaq, B., Haggerty, T., & Brownstein, H. H. (2021). Tweeting Stigma: An Exploration of Twitter Discourse Regarding Medications Used for Both Opioid Use Disorder and Chronic Pain. Journal of Drug Issues, 51(2), 340–357.

Davenport, S., Weaver, A. and Caverly, M. (October 2019). Economic Impact of Non-Medical Opioid Use in the United States. Society of Actuaries. www.soa.org/globalassets/assets/files/resources/research-report/2019/econ-impact-non-medical-opioid-use.pdf.

Engel, L. B., Bright, S. J., Barratt, M. J., & Allen, M. M. (2020). Positive drug stories: possibilities for agency and positive subjectivity for harm reduction. Addiction Research & Theory, 29(5), 363–371

Graham, S. S. (2021). Misinformation Inoculation and Literacy Support Tweetorials on COVID-19. Journal of Business and Technical Communication, 35(1), 7–14.

Hammarlund, R., Crapanzano, K. A., Luce, L., Mulligan, L., & Ward, K. M. (2018). Review of the effects of self-stigma and perceived social stigma on the treatment-seeking decisions of individuals with drug-and alcohol-use disorders. Substance abuse and rehabilitation, 9, 115.

Heley, Kathryn, Alene Kennedy-Hendricks, Jeff Niederdeppe, and Colleen L. Barry. “Reducing health-related stigma through narrative messages.” Health communication (2019).

Ho, J., Tumkaya, T., Aryal, S., Choi, H., & Claridge-Chang, A. (2019). Moving beyond P values: data analysis with estimation graphics. Nature methods, 16(7), 565–566.

Holland, K. M., Jones, C., Vivolo-Kantor, A. M., Idaikkadar, N., Zwald, M., Hoots, B., … & Houry, D. (2021). Trends in US emergency department visits for mental health, overdose, and violence outcomes before and during the COVID-19 pandemic. JAMA psychiatry, 78(4), 372–379.

Howard, H. (2015). Reducing stigma: Lessons from opioid-dependent women. Journal of Social Work Practice in the Addictions, 15(4), 418–438.

Hu-Undark, J. Inside the controversial rise of a top Twitter COVID-19 influencer, FastCompany, https://www.fastcompany.com/90581545/eric-feigl-ding-covid-19-twitter.

Kantor, D., Bright, J. R., & Burtchell, J. (2018). Perspectives from the patient and the healthcare professional in multiple sclerosis: social media and participatory medicine. Neurology and therapy, 7(1), 37–49.

Kelly, J. F., & Westerhoff, C. M. (2010). Does it matter how we refer to individuals with substance-related conditions? A randomized study of two commonly used terms. International Journal of Drug Policy, 21(3), 202–207.

Kelly, J. F., Wakeman, S. E., & Saitz, R. (2015). Stop talking ‘dirty’: clinicians, language, and quality of care for the leading cause of preventable death in the United States. The American journal of medicine, 128(1), 8–9.

Kennedy-Hendricks, A., McGinty, E. E., & Barry, C. L. (2016). Effects of competing narratives on public perceptions of opioid pain reliever addiction during pregnancy. Journal of Health Politics, Policy and Law, 41(5), 873–916.

Kuhn, L., Davidson, L. L., & Durkin, M. S. (1994). Use of Poisson regression and time series analysis for detecting changes over time in rates of child injury following a prevention program. American journal of epidemiology, 140(10), 943–955.

Lavack, A. (2007). Using social marketing to de-stigmatize addictions: A review. Addiction Research & Theory, 15(5), 479–492.

Linas, B. P., Savinkina, A., Barbosa, C., Mueller, P. P., Cerdá, M., Keyes, K., & Chhatwal, J. (2021). A clash of epidemics: Impact of the COVID-19 pandemic response on opioid overdose. Journal of Substance Abuse Treatment, 120, 108158.

Luo, F., Li, M., & Florence, C. (2021). State-level economic costs of opioid use disorder and fatal opioid overdose—United States, 2017. Morbidity and Mortality Weekly Report, 70(15), 541.

Luoma, J. B. (2010). Substance use stigma as a barrier to treatment and recovery. In Addiction medicine (pp. 1195–1215). Springer, New York, NY.

Mattson, C. L., Tanz, L. J., Quinn, K., Kariisa, M., Patel, P., & Davis, N. L. (2021). Trends and geographic patterns in drug and synthetic opioid overdose deaths— United States, 2013–2019. Morbidity and Mortality Weekly Report, 70(6), 202.

McGinty, E. E., & Barry, C. L. (2020). Stigma reduction to combat the addiction crisis—developing an evidence base. New England Journal of Medicine, 382(14), 1291–1292.

McGinty, E. E., Goldman, H. H., Pescosolido, B., & Barry, C. L. (2015). Portraying mental illness and drug addiction as treatable health conditions: effects of a randomized experiment on stigma and discrimination. Social Science & Medicine, 126, 73–85.

McGinty, E. E., Kennedy-Hendricks, A., & Barry, C. L. (2019). Stigma of addiction in the media. In The stigma of addiction (pp. 201–214). Springer, Cham.

National Institute on Drug Abuse (NIDA). (July 2021). Words Matter - Terms to Use and Avoid When Talking About Addiction. https://www.drugabuse.gov/nidamed-medical-health-professionals/health-professions-education/words-matter-terms-to-use-avoid-when-talking-about-addiction.

Neporent, L. (May 12, 2020). Coronavirus social: Twitter hustles to verify physicians. Medscape, https://www.medscape.com/viewarticle/930360.

Pavlov, I. P. (1927). Conditioned reflexes: An investigation of the physiological activity of the cerebral cortex. Oxford Univ. Press.

Saloner, B., McGinty, E. E., Beletsky, L., Bluthenthal, R., Beyrer, C., Botticelli, M., & Sherman, S. G. (2018). A public health strategy for the opioid crisis. Public Health Reports, 133(1_suppl), 24S–34S.

Signorini, D. F. (1991). Sample size for Poisson regression. Biometrika, 78(2), 446–450.

Skinner, B. F. (1938). The behavior of organisms: An experimental analysis. New York, NY: Appleton-Century.

Stangor, C., & Walinga, J. (2014). Introduction to Psychology. BCcampus. https://opentextbc.ca/introductiontopsychology/

Stone, E. M., Kennedy-Hendricks, A., Barry, C. L., Bachhuber, M. A., & McGinty, E. E. (2021). The role of stigma in US primary care physicians’ treatment of opioid use disorder. Drug and alcohol dependence, 221, 108627.

Tornes, A., & Trujillo, L. (January 26, 2021). Enabling the future of academic research with the Twitter API, Twitter Developer Blog, https://blog.twitter.com/developer/en_us/topics/tools/2021/enabling-the-future-of-academic-research-with-the-twitter-api.

Thorndike, E. L. (1905). The elements of psychology. New York, NY: A. G. Seiler.

Tsai, A. C., Kiang, M. V., Barnett, M. L., Beletsky, L., Keyes, K. M., McGinty, E. E., … & Venkataramani, A. S. (2019). Stigma as a fundamental hindrance to the United States opioid overdose crisis response. PLoS medicine, 16(11), e1002969.

Tükel, S. (2020). Motor learning. In S. Angin & I. E. Simsek (eds.), Comparative Kinesiology of the Human Body: Normal and Pathological Conditions (pp. 453–466). Cambridge, MA. Academic Press.

Wakeman, S. E., & Rich, J. D. (2018). Barriers to medications for addiction treatment: How stigma kills. Substance use & misuse, 53(2), 330–333.

Watson, J. B. (1924). Behaviorism. New York, NY: People’s Institute Publishing Company.

